# Regular Khat Use and Its Association with Poverty: A Cross-sectional Study in Ethiopia

**DOI:** 10.64898/2026.07.25.26358415

**Authors:** Awoke Mihretu, Abebaw Fekadu, Azeb Asaminew Alemu, Solomon Teferra, Kassahun Habtamu, Wubalem Fekadu

## Abstract

**Background:** Khat use is widespread in Ethiopia and has increasingly raised economic concerns. Regular khat use may negatively affect productivity, household income, and social functioning. However, limited empirical research has examined its association with poverty. This study, therefore, aimed to examine the association between regular khat use and household food insecurity and wealth status in Ethiopia.

**Methods:** A cross-sectional survey was conducted in the East Gurage Zone of the Central Ethiopia Regional State. 924 individuals randomly selected from the Addis Ababa University Health and Demographic Survey (HDS) registry participated in the study. Regular khat use was defined operationally as chewing khat 3 or more days per week in the past month. Household wealth status was assessed using wealth index indicators, while household food insecurity was measured using the Household Food Insecurity Access Scale (HFIAS). Univariate and multivariable logistic regression analyses were performed to examine the association of regular khat use with poverty-related outcomes (i.e. household food insecurity and wealth status), after adjusting for potential confounders.

**Results:** The proportion of participants who regularly used khat was 57.3%. A higher proportion of regular khat users were classified in the low-wealth category than non-users (52.5% vs 34.9%). Household food insecurity was also higher among khat users than non-users (64.4% vs 35.6%). Regular khat use was significantly associated with low wealth status (AOR = 1.54; 95% CI: 1.11-2.13) and household food insecurity (AOR = 1.98, 95% CI: 1.39,2.83), after adjusting for potential confounders.

**Conclusion:** We found that regular khat use was significantly associated with poverty-related outcomes, particularly household food insecurity. Overall, people who were classified as regular khat users were more likely to experience food insecurity and socioeconomic disadvantages than non-users. Our findings highlight the need for integrated interventions to reduce poverty and improve household food security through coordinated socioeconomic support and targeted strategies to curb regular khat use.

## Background

Khat use is widespread in Ethiopia, and it plays an important role in the social and cultural life of communities [1, 2]. It has also increasingly raised concerns regarding its broader social and economic impact [3]. Khat use has been widely associated with adverse socioeconomic consequences, particularly affecting users’ productivity, income, and overall household economic stability [3]. Evidence suggests that regular khat consumption is associated with substantial time lost from productive activities, with users spending considerable hours per month on khat-related activities[2]. In addition, a significant proportion of household income is likely allocated to khat use. Some estimates suggest that up to 30% of household income may be spent on khat ceremonies[4]. These patterns are often accompanied by reduced work performance, including leaving work early, returning late from breaks, and diminished occupational engagement. Over time, such behaviours contribute to excessive expenditure to khat, debt accumulation, and limited savings, thereby increasing the risk of downward socioeconomic mobility and poverty among users[4].

At a broader societal level, where economic vulnerability is already high, such as in Ethiopia, regular khat use may further exacerbate household and structural poverty through reduced productivity and increased financial strain[3]. The financial burden of sustaining khat consumption, combined with lost working hours during khat sessions, further contributes to household instability, marital breakdown, and divorce[5].

In Ethiopia, these concerns are particularly salient given the growing reliance on khat as a major cash crop. Khat is currently one of the country’s most valuable cash crops, generating an estimated 300 million USD annually[6]. However, its expansion has also driven agricultural restructuring, with farmers increasingly replacing cereal crops with khat cultivation in several regions. While this shift may yield short-term economic returns for khat-producing farmers, there is limited empirical evidence on its long-term implications for household food security, agricultural sustainability, and rural poverty reduction. This highlights the need for a rigorous study of the impact of khat use on household-level wealth status and food security, as well as on the nation’s economy.

Many previous studies have small sample sizes, focus on specific groups, or are qualitative; therefore, additional evidence from a larger sample and a community-based study is warranted. This study aimed to examine the association between regular khat use and household food insecurity and low wealth status in the East Gurage zone of the Central Ethiopia Regional State.

## Methods

### Study design

A cross-sectional survey was conducted as part of a broader cohort study to examine the long-term multidimensional harms of khat use. The study reported in this paper examined the association between regular khat use and poverty, as indicated by wealth status and household food insecurity.

### Study setting

The study was conducted in the East Gurage Zone of the Central Ethiopia region, which includes the study sites, Butajira Town Administration and adjacent rural districts. The area is predominantly agrarian, with peasant farming the main livelihood in rural areas and petty trading common in urban areas. The zone is widely recognised for khat production and consumption and serves as a major supplier to the domestic market in the Southern and Central part of Ethiopia. The agroecological conditions are highly suitable for khat cultivation, contributing to its widespread availability and use in the region. The study site was selected because it is an established research site of Addis Ababa University, particularly the College of Health Sciences.

### Study population and eligibility criteria

The study population consisted of permanent residents aged 18 years and above living in the study area. Individuals were eligible for inclusion if they were between 18 and 65 years of age and had been permanent residents of the selected sub-districts for at least 2 years.

### Sample size determination

Sample size was calculated for multiple outcomes as part of the broader cohort research project, assuming a 1:1 ratio of exposed to non-exposed participants. Using STATCALC with 80% power, a two-sided 95% confidence level, a 50% prevalence of khat use, and an anticipated 15% loss to follow-up, the target sample size was 930 participants (8).

### Participant selection

Participants were identified through the Addis Ababa University Health and Demographic Survey (HDS) registry, which assigns each individual a unique 10-digit identification number that incorporates district, household, and individual identifiers. A comprehensive list of residents was extracted from the registry and stratified by age and sex. Within each eligible household, one individual was randomly selected from the registry to participate.

## Measures

### Household food insecurity (HFIAS)

Household food insecurity was assessed using the Household Food Insecurity Access Scale (HFIAS), a nine-item instrument with a polytomous response format [7]. The scale measures the access dimension of food insecurity, including anxiety or uncertainty about food supply, inadequate food quality in terms of variety and preferences, and insufficient food intake. The HFIAS has been widely used in both rural and urban settings in Ethiopia, with evidence supporting its acceptable psychometric properties, acceptable construct validity, effective distinction between different levels of access-related food deprivation among households and good internal consistency[8, 9]. We have used an adapted version of the tool, which could improve contextual understanding and respondent engagement [10].

Each HFIAS item is scored by frequency of occurrence, ranging from “rarely” to “often,” yielding a total score between 0 and 27, with higher scores indicating greater household food insecurity. Although the instrument classifies households into four categories—food secure, mildly food insecure, moderately food insecure, and severely food insecure—based on the severity of reported conditions, for the current analysis the categories were collapsed into two to enhance the interpretability of the regression model we fitted [7]. Accordingly, food-secure and mildly food-insecure households were combined into a single category, while moderately and severely food-insecure households were grouped as food-insecure. The original HFIAS guidance by Coates et al. (2007) allows for flexible use of the scale, including binary classification of food security versus food insecurity depending on analytic objectives.

### Wealth Status

Wealth status determination followed the standard Demographic and Health Survey (DHS) approach, using principal component analysis of household asset variables, as described by The DHS Program and consistent with the Ethiopia DHS survey methodology[11]. Household assets and housing characteristics provided wealth scores after applying principal component analysis (PCA). Household asset variables (e.g., cooking fuel type, toilet facility, floor material, water source, and selected durable assets) were first recoded into binary variables (0 = not owned/absence, 1 = owned/presence). For categorical variables with a single response (e.g., type of cooking fuel), these were similarly transformed into a set of binary indicators using dummy coding.

Principal component analysis was then performed on the standardised binary asset variables to generate a composite wealth index score. The first principal component (FAC1_1), which explained the largest proportion of variance in the asset data, was retained as the household wealth index score[12]. To ensure comparability with standard population distributions, national reference cut-off points were obtained from the DHS Program dataset documentation (Household Recode files). The 20th, 40th, 60th, and 80th percentile cut-points were derived from the national wealth index distribution and applied to the study data to construct wealth quintiles.

Accordingly, households were initially classified into five wealth quintiles: Q1 (poorest), Q2 (poorer), Q3 (middle), Q4 (richer), and Q5 (richest). For analysis purposes, these quintiles were further collapsed into two categories: the lower three quintiles (Q1–Q3) were classified as “low wealth status,” and the upper two quintiles (Q4–Q5) as “high wealth status,” for ease of interpretation in regression analysis.

### Regular khat use

Khat use was assessed using a structured questionnaire that captured different patterns of use, including current khat use and frequency of use[2]. For the present study, regular khat use was considered the primary exposure variable. Regular khat use was operationally defined as chewing khat on three or more days per week during the past month [2].This definition was used to distinguish habitual users from occasional or infrequent users and to better capture sustained patterns of khat consumption that may have socioeconomic implications.

### Assessment of other covariates

Other covariates in the current study were problematic alcohol use, stressful life events, smoking status, and sociodemographic characteristics. Problematic alcohol use was assessed using the Alcohol Use Disorders Identification Test (AUDIT), a 10-item instrument that evaluates alcohol consumption patterns, drinking frequency, quantity of alcohol intake, and alcohol-related adverse consequences over the past 12 months. A score of 8 or higher on the AUDIT is considered indicative of high-risk alcohol use[13]. Stressful life events were measured using the List of Threatening Experiences (LTE), which assesses major adverse events such as the loss of important relationships, the death of close family members or friends, financial crises, and imprisonment[14]. The LTE consists of dichotomous items with “yes” or “no” response options. A cut-off score of 1 or higher was considered indicative of “experience of LTE.” Smoking status was assessed with a single-item question about cigarette smoking over the past 30 days, with responses categorised as yes or no. Additional covariates, including age, sex, educational status, and marital status, were assessed using a structured questionnaire.

### Data collection procedure

Data were collected through a house-to-house survey conducted by trained data collectors. Structured questionnaires were administered to collect data. Data were captured using REDCap electronic data capture tools. Field activities were closely supervised throughout the data collection period to ensure data quality, completeness, and consistency.

### Data quality assurance

A comprehensive three-day training was provided for supervisors and data collectors focusing on study objectives and methods, study instruments, data collection procedures, and ethical issues. The training also included detailed reviews of the questionnaires, role plays, and pilot testing, followed by debriefing sessions to ensure consistency and understanding. Data collection involved at least two rounds of household visits, and Global Positioning System (GPS) coordinates were recorded for all households to support accurate tracking and follow-up. Continuous field supervision was implemented throughout the data collection period to ensure adherence to protocols.

### Data Analysis

Data were analysed using descriptive and inferential statistical methods. Descriptive statistics, including mean, median, interquartile range, frequencies, and percentages, summarised participants’ sociodemographic and clinical characteristics across categories of wealth status and food security. For ease of interpretation of the regression analyses, the outcome variables (i.e., wealth status and food security) were recoded into binary variables. Similarly, the main exposure variable, regular khat use, which was initially assessed across multiple frequency categories ranging from monthly to daily, was dichotomised as chewing khat on three or more days per week during the past month versus less than three days per week (yes/no). Univariate logistic regression analyses were first conducted to examine the crude associations between regular khat use and each outcome variable, as well as the covariates. Variables considered theoretically relevant or associated with the outcomes in the univariate analysis were subsequently entered into the multivariable logistic regression models to estimate adjusted odds ratios (AORs) with corresponding 95% intervals (CIs), while controlling for potential confounding factors. Statistical significance was determined using a p-value of less than 0.05.

### Ethical Considerations

Ethical clearance was obtained from the Addis Ababa University College of Health Sciences Institutional Review Board (IRB) (Ref: 023/24/Psy). Written informed consent was obtained from all participants prior to data collection. Detailed information about the study was provided to participants before they decided to participate. All interviews were conducted in a private setting in the participants’ homes or a private public area. There was no personally identifiable information included in the dataset. Data were stored securely and were accessible only to the research team.

## Results

### Socio-demographic and clinical characteristics of participants

The study included 924 participants with a mean age of 37.2 years (SD ≈ 10.8). The majority were men (65%) and married (80.2%). More than half of the participants (57.3%) were regular khat users, while 42.7% were non-users. Regarding socioeconomic characteristics, the majority of participants were from food-insecure households (668, 72.3%), while 256 (27.7%) participants were from food-secure households. In terms of wealth status, 414 (44.9%) participants were classified as having low wealth status, whereas 507 (55.1%) were categorised as having average and above wealth status (Table 1).

**Table 1:** Demographic, socioeconomic and clinical characteristics of study participants (N=924)

| Characteristics | Category | n (%) |
| --- | --- | --- |
| Regular khat use | No | 393 (42.7) |
|  | Yes | 528 (57.3) |
| Age (in years) | Mean ± SD | 37.4 ± 10.9 |
| Marital status | Single | 127 (13.8) |
|  | Married | 739 (80.2) |
|  | Divorced/Widowed | 56 (6.1) |
| Residence | Rural | 530 (57.6) |
|  | Urban | 390 (42.4) |
| Sex | Male | 596 (64.8) |
|  | Female | 324 (35.2) |
| Educational status | Cannot read/write | 160 (17.4) |
|  | Read and write only | 56 (6.1) |
|  | Primary school | 396 (43.0) |
|  | Secondary school | 233 (25.3) |
|  | College/University | 77 (8.4) |
| Alcohol use (AUDIT) | Low-risk alcohol use | 883 (95.8) |
|  | High-risk alcohol use | 39 (4.2) |
| Smoking status | Nonsmoker | 879 (95.8) |
|  | Smoker | 39 (4.2) |
| Stressful life events | No | 456 (49.5) |
|  | Yes | 466 (50.5) |
| Household food security | Food insecure | 668 (72.3) |
|  | Food secure | 256 (27.7) |
| Wealth status | Low wealth | 414 (44.9) |
|  | High wealth | 507 (55.1) |

#### Khat Use and Wealth Status

Regular khat use was significantly associated with lower wealth status in both the crude (COR = 2.08, CI 1.59,2.70) and adjusted (AOR = 1.54, CI 1.11,2.13) models. Increasing age was significantly associated with average and above wealth status in the adjusted model (COR= 0.99, CI 0.98–1.00 and AOR = 0.96, CI 0.94,0.98). Stressful life events were also significantly associated with lower wealth status (COR= 2.56, CI 1.96–3.33 and AOR = 2.38, CI 1.82,3.23). Divorced or widowed individuals had higher odds of poor wealth status than singles in the adjusted model (COR= 1.89, 0.99–3.57 and AOR = 2.50, CI 1.15,5.56) (Table 2).

**Table 2:**
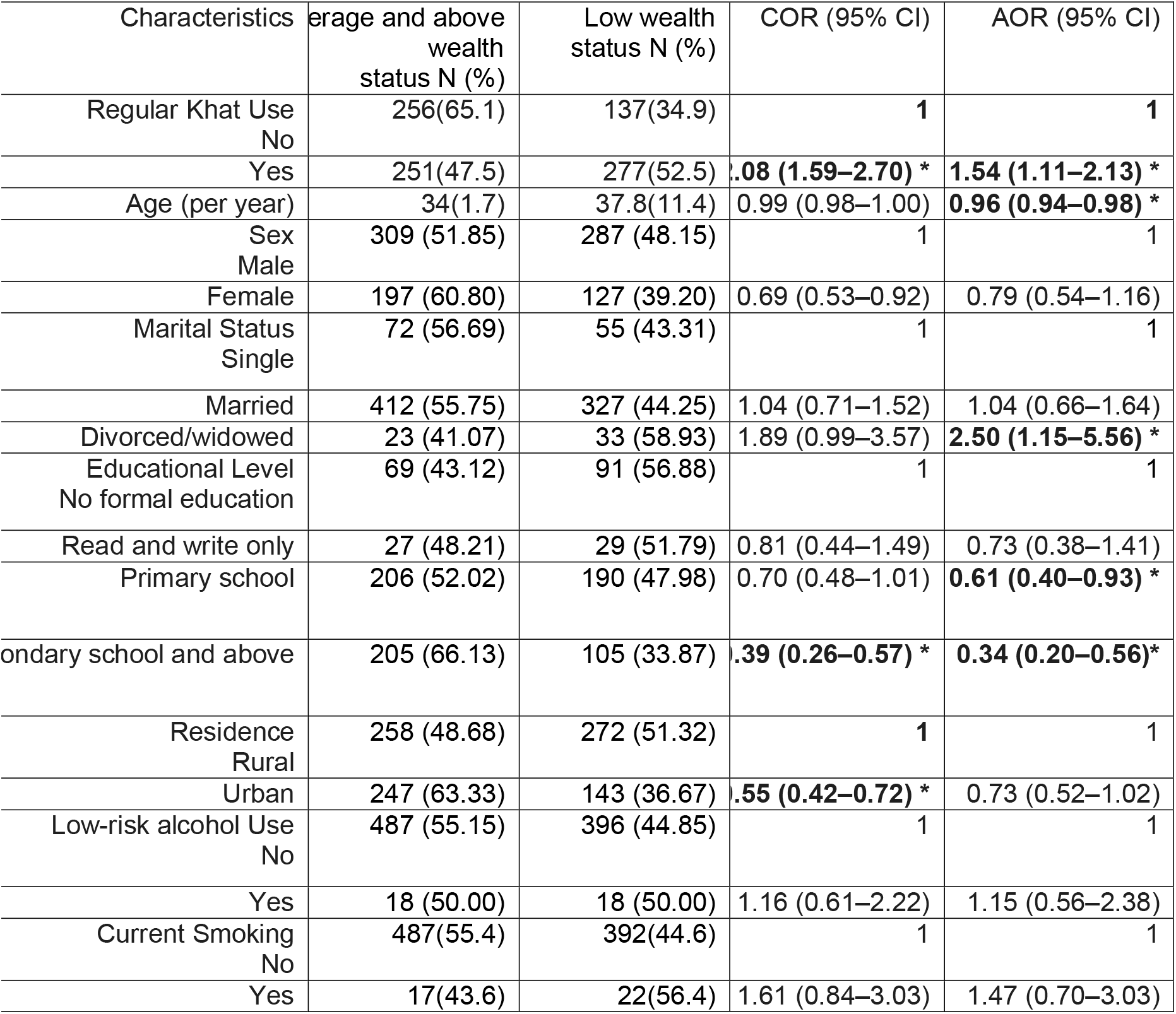
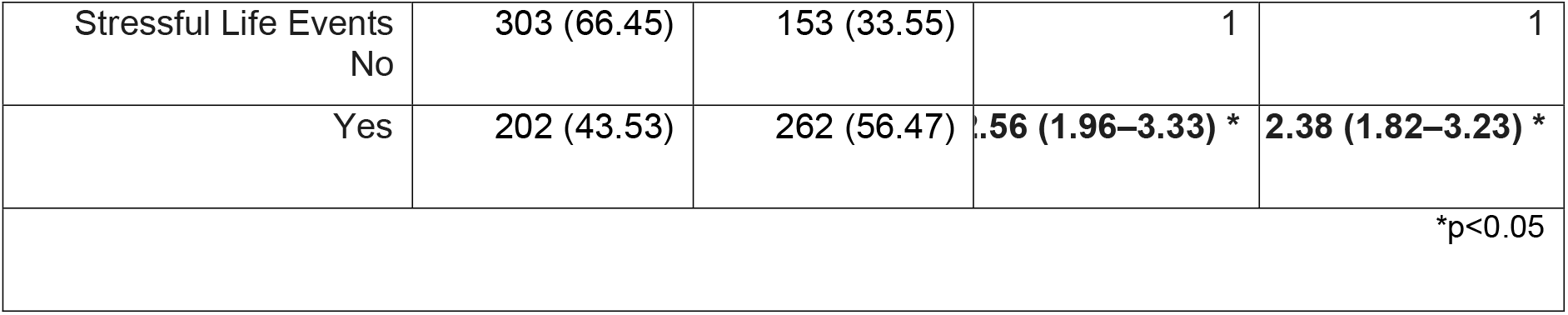
Crude and adjusted odds ratios for the association between wealth status and regular khat use (N=924)

#### Khat Use and Household Food Insecurity

Regular khat use was significantly associated with higher odds of household food insecurity, both in the unadjusted model (COR = 2.82, CI 2.09–3.79 and after adjusting for confounding variables (AOR = 1.98, CI: 1.39–2.83). Rural residence was associated with lower odds of household food insecurity (COR = 0.37, CI 0.27–0.49; AOR = 0.45, CI 0.31–0.64). Experiencing stressful life events also increased the odds of household food insecurity (COR = 1.80, CI 1.35– 2.42; AOR = 1.58, CI 1.15–2.16) (Table 3).

**Table 3:**
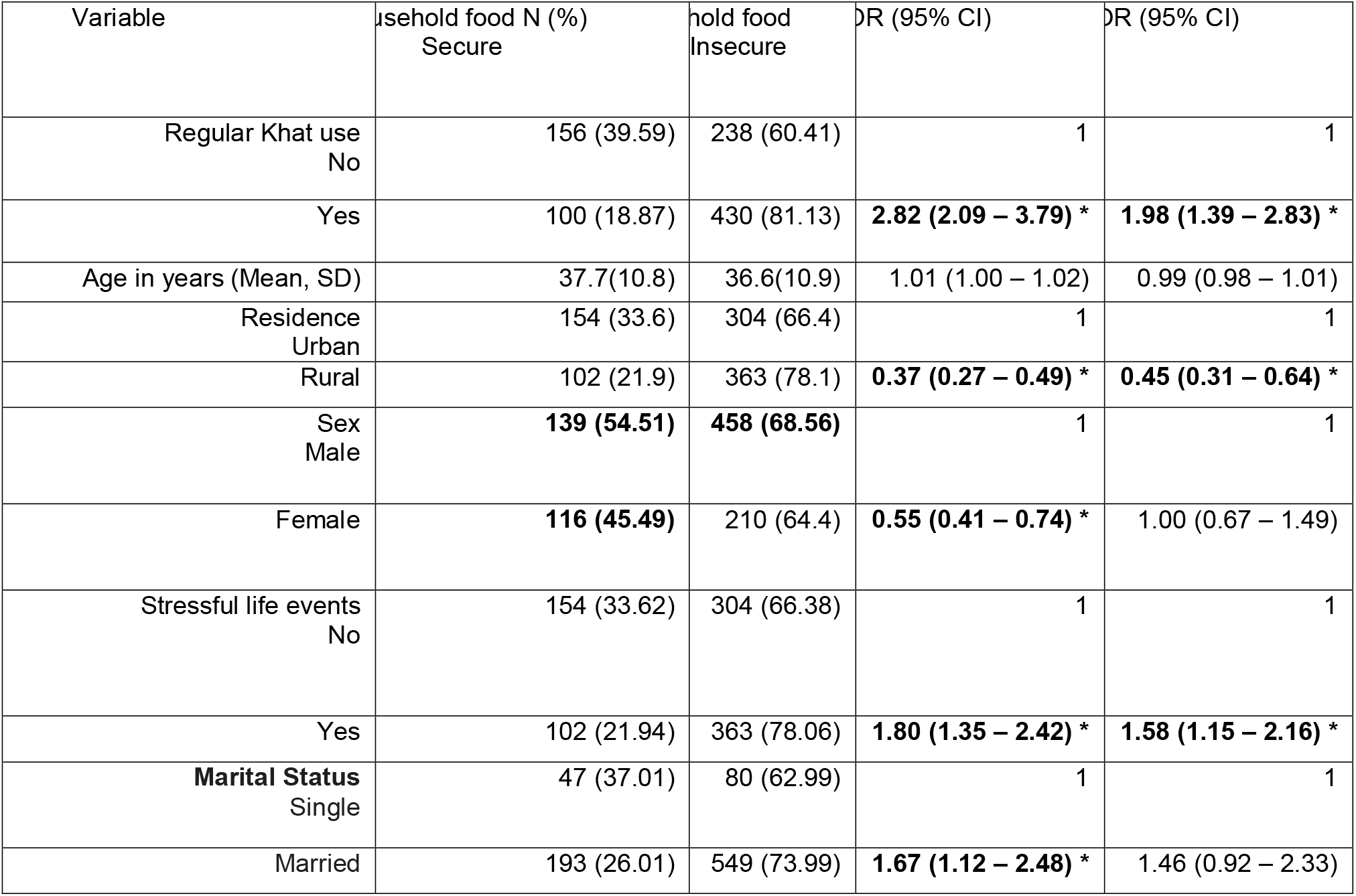
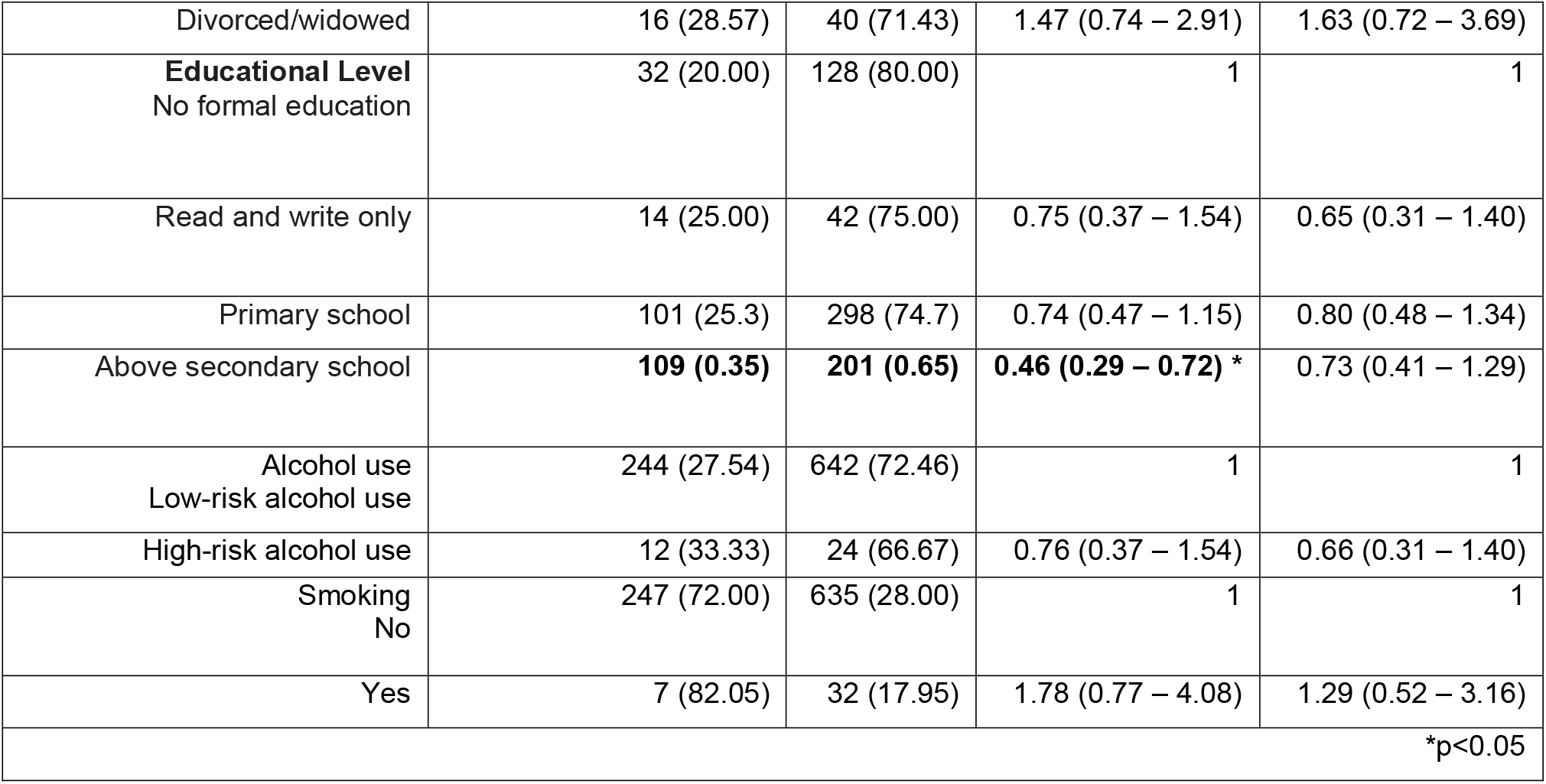
Crude and adjusted odds ratios for the association between household food insecurity and regular khat use (N=924)

## Discussion

This study provides evidence on the association between regular khat use and poverty, as measured by household food insecurity and wealth status. The proportion of individuals in the lowest wealth quartiles in the study population was approximately 45%, higher than the national estimate of 33% [15], suggesting a possible concentration of economic disadvantage within this population.

The estimated poverty in this study is consistent with findings from other recent reports. According to the World Bank’s World Development Indicators, Ethiopia’s predicted poverty headcount ratio at the international poverty line of US$3.00 per person per day (2021 PPP) was 43.3% in 2025[15]. In 2024, 61% of Ethiopians experienced moderate or high lived poverty, as measured by the Afrobarometer Lived Poverty Index (LPI) [16]. According to the Global Multidimensional Poverty Index (MPI) 2024, 72% of Ethiopians were multidimensionally poor[16].

The prevalence of household food insecurity among regular khat users in this study is substantial, indicating a significant burden of deprivation. A previous study in a neighbouring region of Ethiopia, using a similar measure, reported that 52.9% of the general population experienced household food insecurity[17]. The observed level of food insecurity is also broadly consistent with findings among populations with severe mental illness, where severe food insecurity has been reported at approximately 32.5% when using narrower definitions that exclude moderate insecurity (15).

In this study, regular khat use was significantly associated with adverse socioeconomic outcomes, including household food insecurity and lower wealth status. Participants who reported regular khat use had higher odds of experiencing household food insecurity compared with non-users, even after controlling for potential confounding factors. Similarly, regular khat use was independently associated with lower wealth status, suggesting that frequent khat consumption may be linked to socioeconomic disadvantages.

The relationship between khat use and socioeconomic disadvantage is likely to be bidirectional. On the one hand, regular khat use may contribute to poverty and food insecurity through the diversion of household income, reduced productivity, loss of working hours, and increased khat-related expenditure. On the other hand, individuals experiencing poverty and socioeconomic instability may be more vulnerable to initiating or maintaining khat use as a coping strategy within constrained social and economic environments [18]. This bidirectional relationship highlights the complexity of disentangling cause and effect in cross-sectional data.

The findings of this study contrast with some previous reports that have suggested positive associations between khat use and social or economic functioning in certain contexts[19]. While it is acknowledged that khat production and trade can provide livelihoods and income for some groups, particularly producers and traders, the economic benefits appear to be unevenly distributed [20]. A substantial proportion of khat users may instead experience net economic loss due to consumption-related expenditures and associated functional impairment. This reinforces the importance of distinguishing between khat’s economic role as a commodity and its effects on users at the individual and household levels.

The findings of the present study add to the growing empirical evidence that regular khat use is associated with adverse economic outcomes. Prior studies have documented several mechanisms through which khat use may undermine household welfare, including reduced productivity, diversion of income away from essential needs, increased health-related expenditures, interpersonal conflict, marital disruption, and nutritional consequences[3, 4]. For example, evidence from Yemen and Djibouti indicates that some individuals allocate up to one third of their income to khat consumption, illustrating the potential scale of economic diversion [21]. Similarly, studies from Somalia have reported family dysfunction and behavioural changes associated with chronic use, further highlighting the broader social consequences [22, 23].

Overall, the findings of the present study have important economic and public health implications, particularly for household food insecurity. At the population level, regular khat use may contribute to preventable economic losses through reduced productivity, diversion of household income, and increased financial strain. Addressing regular khat use through effective policy, prevention, and intervention strategies may improve household economic stability, enhance participation in productive labour, and reduce the broader social and economic burdens associated with regular khat use.

## Conclusion

The study found that a higher proportion of individuals who regularly use khat were in the lowest wealth quartile and experienced household food insecurity. Regular khat use was significantly associated with poverty-related outcomes, including household food insecurity and low wealth status. Integrated interventions that combine behavioral counselling and community-based prevention strategies with socioeconomic support measures, such as livelihood strengthening, poverty reduction initiatives, and household food security programs, are therefore warranted to address regular khat use alongside the underlying socioeconomic vulnerabilities.

## Data Availability

All are included in the manuscript.

## Authors’ contributions

All authors contributed to the study’s design, and A.M. led the study. A.M. and W.F. contributed to data collection and analysis. A.M. drafted the manuscript under the supervision of W.F. and K.H. All authors made critical intellectual contributions, reviewed and approved the final manuscript, and agreed to its submission for publication.

## Acknowledgments

The authors acknowledge the contributions of the study participants. We also acknowledge Addis Ababa University for funding support.

## Statements and Declarations

Nothing to declare.

## Ethics Approval

Ethical clearance was obtained from the Addis Ababa University College of Health Sciences Institutional Review Board (IRB) (Ref: 023/24/Psy). Written informed consent was obtained from all participants prior to data collection. For participants who could not read or write, a neutral witness was present during the consent process and signed a witness statement confirming that the study information and consent form had been explained and understood by the participant, and that consent was provided voluntarily. Detailed information about the study was provided to participants before they decided to participate. All interviews were conducted in a private setting, either in participants’ homes or in private public areas. No personally identifiable information was included in the dataset. Data were stored securely and were accessible only to the research team.

## Consent for publication

Not applicable.

## Data availability

All relevant materials and data supporting the findings of this study are included within the manuscript. Additional documents may be made available upon reasonable request.

## Declaration of conflicting interest

The authors declared no potential conflicts of interest with respect to the research, authorship, and/or publication of this article.

## Funding

This study was funded by Addis Ababa University through its Thematic Research Project Fund Scheme.

## Notes

### Competing Interest Statement

The authors have declared no competing interest.

### Author Declarations

Ethical clearance was obtained from the Addis Ababa University College of Health Sciences Institutional Review Board (IRB) (Ref: 023/24/Psy)

